# Exploring the help-seeking journeys for Long Covid from a health inequalities perspective: a qualitative study in England

**DOI:** 10.1101/2024.12.12.24318614

**Authors:** Donna Clutterbuck, Mel Ramasawmy, Marija Pantelic, Gail Allsopp, Mark Gabbay, Jasmine Hayer, Melissa Heightman, Yi Mu, David Sunkersing, Dan Wootton, Amitava Banerjee, Nisreen A Alwan, STIMULATE-ICP Consortium

**Affiliations:** School of Primary Care, Population Sciences and Medical Education, University of Southampton, Southampton, UK; Institute of Health Informatics, University College London, London, UK; NIHR Applied Research Collaboration Wessex, Southampton, UK; Brighton and Sussex Medical School, University of Sussex, Falmer, UK; Department of Social Policy and Intervention, University of Oxford, Oxford, UK; Royal College of General Practitioners, London, UK; School of Medicine, University of Nottingham, Nottingham, UK; Department of Primary Care and Mental Health, University of Liverpool, Liverpool, UK; NIHR Applied Research Collaboration North West Coast, Liverpool, UK; PPIE co-applicant for STIMULATE-ICP; University College London Hospitals NHS Trust, London, UK[]; Clinical Infection, Microbiology and Immunology, University of Liverpool, UK; Liverpool University Hospitals NHS Foundation Trust, Liverpool, UK; University Hospital Southampton NHS Foundation Trust, Southampton, UK

## Abstract

**Background and aim:** Long Covid is a health condition that continues to be challenging in terms of obtaining care and support, even in the fourth year following its emergence. This study, which forms part of the STIMULATE-ICP study in England, explores the barriers and facilitators people with Long Covid face when trying to access care, as well as experiences in relation to stigma, discrimination, and inequitable treatment.

**Methods:** The study was co-designed with people with lived experience of Long Covid. People attending three post-covid services in England were invited to participate by clinic staff. Twenty-three participants were interviewed about their experiences in relation to barriers and facilitators of accessing adequate care, including experiences of being treated unfairly. Interviews were analysed thematically.

**Findings:** Participants experienced difficulties in accessing and receiving appropriate support from primary and secondary care but generally care and support improved once participants were under the care of a Long Covid service. Positive interactions with clinicians who were knowledgeable and supportive helped to foster good patient experiences when accessing Long Covid care. Inequalities in accessing care were reported in the form of experiences of gender and race discrimination. People with previous and existing conditions reported further stigmatisation. Financial barriers to care existed and there were also difficulties faced by those who got infected with COVID-19 early in the pandemic. The impact of Long Covid on mental health was evident, as was the stigma related to mental health and the inadequacy of mental health service provision for people with Long Covid.

Some participants who worked within the National Health Service (NHS) perceived their professional position as a facilitator to accessing Long Covid care. However, some NHS employees also reported the negative impact of Long Covid on their work, the lack of employment support available, mistreatment from colleagues, and dismissal of professional knowledge.

**Conclusion:** Our findings highlight a range of barriers to accessing adequate Long Covid care, with women, ethnic minorities and people with co-occurring conditions experiencing intersectional stigma. We recommend a move towards a healthcare system that is sensitive to intersectional disparities in access to care and is mindful of how stigma can reinforce these inequalities. This would speak to removing barriers to care and foster a more positive experience for people living with Long Covid already disadvantaged by structural and systemic discrimination.

## 1. Introduction

Long Covid is defined by symptoms that continue following a confirmed or probable SARS-CoV2 infection for at least three months and are not explained by any other illness or condition.^1,2^ Symptoms of Long Covid can affect multiple body systems, can be constant, fluctuating or relapsing-remitting, potentially causing impairment of everyday functioning. Symptoms include fatigue, breathlessness, headache, cognitive dysfunction, chest pain, muscle/joint pains, cough, disturbed sleep, and neuropsychiatric symptoms, which impact on the lives of those affected.^1,3,4–6^

Long Covid prevalence estimates vary widely depending on type of assessment, risk factors and vaccination status,^3^ however Office for National Statistics (ONS) estimates suggest that up to March 2024, 2 million people in the United Kingdom reported experiencing Long Covid.^7^ Furthermore, an estimate of around 10% of SARS-CoV2-infected people continuing to experience lasting-symptoms is frequently quoted, with at least 65 million people worldwide estimated to suffer from the condition.^8^ However, adults who have received a COVID-19 vaccine are 50% less likely to experience Long Covid.^9^ NHS England announced the commissioning of dedicated Long Covid services to offer support to people with Long Covid symptoms in England in late 2020.^10,11^ These multidisciplinary services were set up to diagnose people living with Long Covid and treat associated symptoms under the umbrella of one service.^10^ In April 2023, most people who received an initial assessment by a Long Covid service in England were from a White ethnic background (86%) and only 20% resided within an area considered to be in the most deprived 20% of areas in England, according to Index of Multiple Deprivation (IMD).^12,13^

Health inequalities are ‘avoidable, unfair and systematic differences in health between different groups of people’.^14^ Health inequalities include disparities in health between different population groups that can result from variations in access and experiences of treatment and healthcare. Little is known about disadvantaged groups’ experiences of Long Covid services, however research is emerging to highlight experiences of discrimination and dismissal among ethnic minority groups.^15^ As with other long-term conditions, evidence suggests that inequalities may exist in access, referral and representation.^15–17^

Inequalities can be shaped by unfair treatment due to an individual’s demographic characteristics, health, location, access to resources, or combinations of these factors.^14^ Intersectionality refers to ‘the critical insight that race, class, gender, sexuality, ethnicity, nation, ability, and age operate not as unitary, mutually exclusive entities, but as reciprocally constructing phenomena that in turn shape complex social inequalities’.^18^ Intersectionality critical theory conceptualises knowledge as contextual, relational, and reflective of political and economic power.^19^ The concept of intersectionality can be used to frame how various forms of disadvantage interact with each other to shape health outcomes,^20^ including in reference to people living with Long Covid.

Emerging evidence suggests that Long Covid is a highly stigmatised condition.^21–23^ The stigma associated with Long Covid is likely to interact with other dimensions of inequality, such as gender and ethnicity.^24^ People with Long Covid report experiencing overt discrimination (enacted stigma). Many people with Long Covid also expect to be discriminated against (anticipated stigma), and hold self-stereotyping beliefs related to Long Covid (internalised stigma).^21^ These various stigma mechanisms can result in further disadvantage for marginalised groups.^24^ For example, people may be denied access to services (overt discrimination) or they may become reluctant to use them even when available due to fear of being treated unfairly (anticipated stigma). People who internalise stigma and develop a sense of low self-worth might also find it difficult to engage with services, especially if these are not sensitively designed and delivered.

Quantitative Long Covid inequalities research is emerging. Qualitative Long Covid studies which detail patient perspectives suggest that people with Long Covid may experience barriers to obtaining care for symptoms^25^ and sometimes these barriers are exacerbated by preexisting disadvantage.^15,26^ Exploring the lived experiences of people with Long Covid in accessing care requires further investigation to inform meaningful system change. This study was conducted in the context of Long Covid healthcare services in England to understand the difficulties people with Long Covid face when accessing these services, while exploring participants’ intersectional lived experiences in relation to stigma, discrimination, and inequitable treatment in seeking and accessing healthcare.

## 2. Materials and methods

This study forms part of the National Institute for Health and Care Research (NIHR) funded STIMULATE-ICP (Symptoms, Trajectory, Inequalities and Management: Understanding long COVID to Address and Transform Existing Integrated Care Pathways) study (NIHR: COV-LT2-0043). This manuscript contributes to the part of STIMULATE-ICP which explores health inequalities in relation to accessing Long Covid care.^27^ This study focuses on experiences of people with Long Covid who have accessed treatment care and support. Other STIMULATE-ICP research into health inequalities has focused on people with Long Covid who have not yet accessed support.^23^

### 2.1 Study design

The study was open to all adults attending Long Covid services within the STIMULATE-ICP study sites who were willing and able to consent to taking part. Long Covid service staff invited people with Long Covid to take part in-person after patient consultations, via email, through patient portals and though clinic-run social media. A member of the research team also visited one clinic to promote the study. Participants were told that the research team were looking for participants to share their experiences of Long Covid care and their experiences related to inequalities. Those interested were encouraged to contact the research team to receive the study participant information sheet (PIS).

Potential participants were directed to read the PIS and encouraged to ask questions for further information or clarification. If they were happy to participate, the consent form was then sent to potential participants for completion before an interview was scheduled. Recruitment was closed once it was felt a breadth of experience related to inequalities was captured across the three Long Covid services. This sample size is also comparable to other qualitative Long Covid studies that include interviews with people with Long Covid.^22,23,25,26^

Semi-structured interviews were conducted with participants who consented to take part in the study via video call (Zoom or MS Teams) between July 2022 and April 2023. Participants were informed at the beginning that interviews were confidential, that the research team had no clinical contact with their Long Covid service or medical team, and that their participation in the study or refusal to participate would not affect their access to care. They were also given an additional opportunity to ask any questions and were reminded that the interview could be paused or stopped at any point.

The interview topic guide consisted of open questions related to positive and negative experiences of Long Covid care, exploring barriers and facilitators, including experiences of being treated unfairly and/or discrimination, as well as what could be done to improve Long Covid care. Patient and public advisors contributed towards the topic guide design. With participant consent, interviews were digitally recorded using the in-built recording function embedded within the video-call software. Demographic information was collected at the end of the interviews after recording had stopped.

Participants who completed the interviews were offered a shopping voucher as a gesture of thanks. Interviews were fully transcribed by an external transcription company. Transcripts were then checked for accuracy prior to analysis. Two interview participants provided additional information by email. This was included in the analysis.

### 2.2 Analysis

Guided by Braun and Clarke’s approach^28,29^ and aided by NVivo,^30^ thematic analysis was used to analyse interview data. Initial coding was completed by DC using an inductive approach. MR performed additional coding on a sub-sample of transcripts. After initial coding, DC and MR began to delve deeper into barriers that related to dimensions of inequality and other life experiences. These experiences were mapped into a table in Microsoft Excel, with example quotes to illustrate these experiences. This table aided in the identification of themes across the sample. DC, NAA (who had lived experience of Long Covid), MR, and MP then met multiple times to discuss and agree on themes before drafting this paper.

### 2.3 Public and Patient Involvement (PPI)

Patient advisors have been involved in study conceptualisation and funding acquisition, study design and the design of study documentation. STIMULATE-ICP continues to be guided by a team of eleven patient advisors.

## 3. Results

### 3.1 Sample characteristics

Twenty-three interviews were completed with people with Long Covid who had received a referral to a Long Covid service. The majority of participants were female, White British and aged 40-49. Most participants reported co-morbidities and just under half worked within the NHS, including doctors, nurses, allied health professionals, and within mental and public health teams.

**Table 1.**
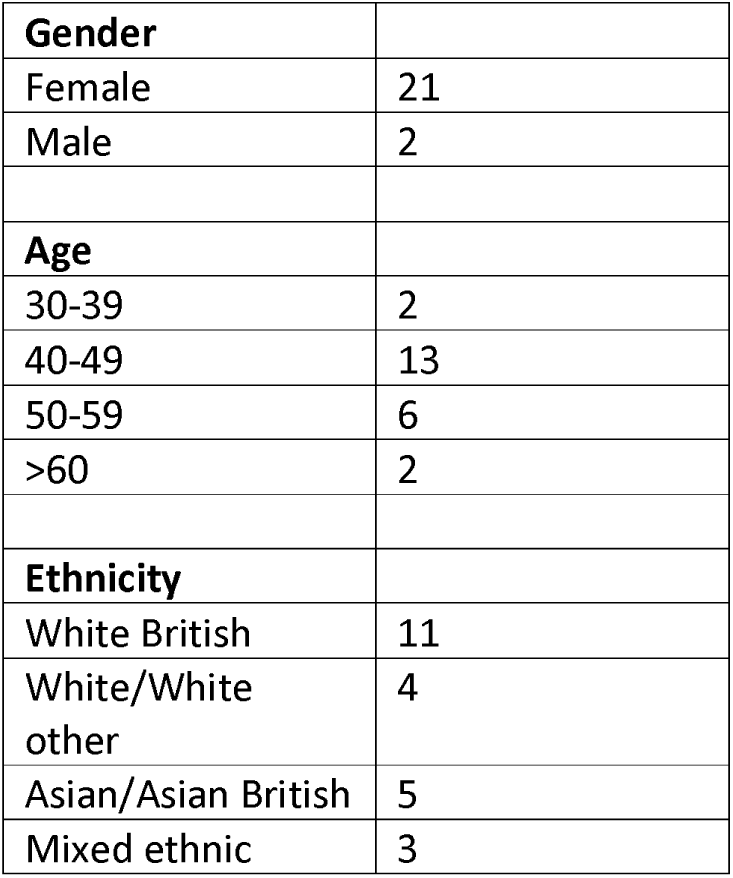
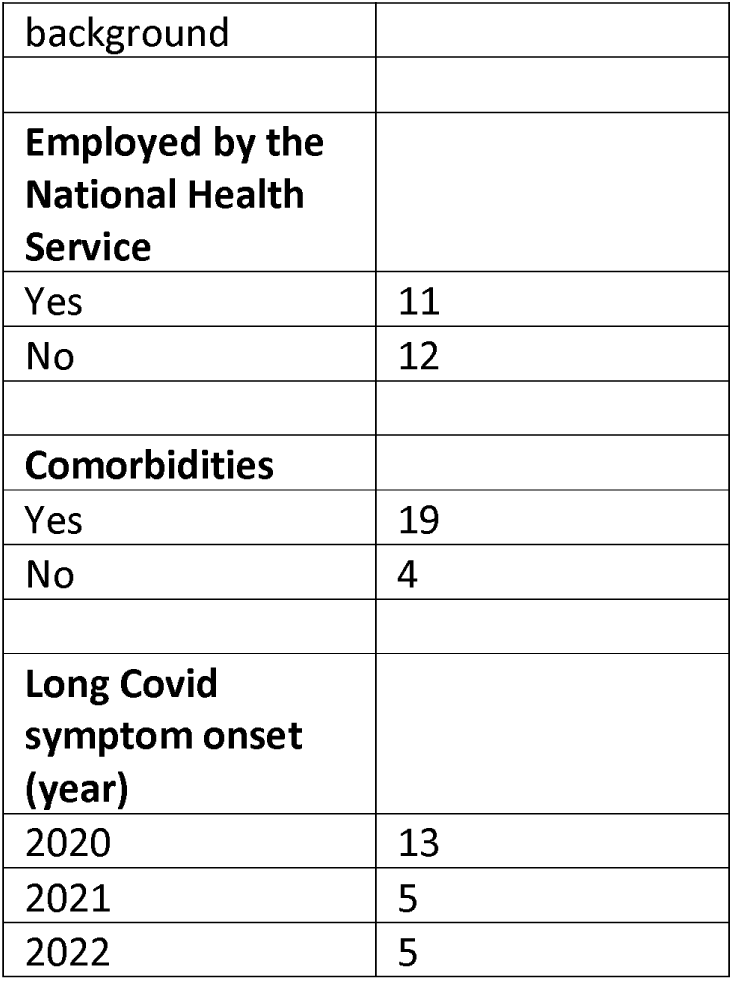
Characteristics of participants.

Participants discussed difficulties and facilitators of accessing and receiving appropriate care from primary care, as well as obtaining a referral, and waiting times to be seen after referrals had been made to specialist secondary care and Long Covid services. Some participants described the period of waiting to be seen as a ‘no man’s land’ or ‘being left to cope alone’. Some participants were still waiting for referrals to be fulfilled at the time of interview. These included referrals for diagnostic scans, respiratory assessment and psychological support.

Participant experiences are discussed in relation to four main themes: 1. Promising avenues for Long Covid care 2. Disparities when seeking access to Long Covid care, 3. Long Covid and mental health, and 4. Experiences of NHS professionals with Long Covid.

Although this study did not to set out to recruit healthcare staff, a significant proportion of this study sample worked within the NHS and some of the findings are unique to this group of participants.

Theme 4 reflects the experiences of NHS workers living with Long Covid, while all other themes discuss experiences across the sample.

### 3.2. Theme 1: Promising avenues for Long Covid care

Every participant described at least one positive experience when attempting to access care for their symptoms that highlighted the differences ‘good’ Long Covid care can make.

#### 3.2.1. Positive experiences with primary care

Many participants described positive experiences when engaging with primary care. Some participants talked about the support they received from their General Practitioner (GP).

> *But my own GP, very good, I didn’t feel fobbed off by her, which I think a lot of other people have done I believe. (Female, 40-49, White British)*
>
> *So there was one who was really good… and would ring me more than once a week to check how I was doing, what my SATs [blood oxygen saturation levels] were, and how I was going. (Female, 40-49, Asian British)*

Additionally, some participants described their GPs as referring them to Long Covid services as soon as they could. Other participants felt supported, but that Long Covid was not originally being suspected by GPs early on, resulting in delays to care.

> *So they took me seriously but…they didn’t think it was COVID at the beginning so it was just many random bad experiences that happened one after the other. (Female, 40-49, White other)*

Similarly, some participants acknowledged the desire to help and support by primary care teams but felt that a lack of knowledge surrounding Long Covid meant GPs have been limited in what support they can give.

> *I see from my own GP the frustration in him that he wishes he could do more. It’s not that he doesn’t want to he just, he doesn’t really know what to do yet so I mean he’s been lovely the way they’ve dealt with it and you know they’ve been good but obviously they’ve got limited experience knowing what to do with it at the minute or things aren’t being filtered through to them or that I don’t know. (Female, 40-49, White)*

#### 3.2.2. Positive experiences and aspirations with clinicians at the Long Covid service

Despite the difficulties which participants experienced when accessing and receiving appropriate care, most, though not all, participants acknowledged that their situations improved once under the care of a Long Covid service. One participant specifically described their first appointment at a Long Covid clinic as *‘a breath of fresh air’*. Others described feelings of being relieved and hopeful. This was often due to their interactions with clinicians working in these services. Participants expressed how they appreciated helpful, supportive clinicians, who believed and validated their experiences.

> *I have liked the conversations with the clinicians. They are very open…I am sure they shouldn’t be giving me as much time as they give me but they give me the time. They reassure me that I am not mad. They do lots of tests. They take me seriously. They refer me for all of my symptoms and take everything seriously. They are brilliant. (Female, 40-49, Asian British)*
>
> *I was…thankful… they recognised my symptoms. They didn’t dismiss me. They knew what was going on, and the head doctor immediately said, “I see hundreds of you. I have seen hundreds of people like you.” So, that was a relief. (Female, 40-49, White)*

Participants welcomed clinicians’ honesty about the extent of their Long Covid knowledge.

> *For someone just to say, “Look we just don’t understand it but we totally get it but we just don’t understand it but we’re doing this and doing that,” which is great which is really helpful and really reassuring. (Female, 40-49, White British)*

Some participants also valued clinicians sharing their own lived experience of Long Covid, creating a sense of understanding.

> *I mean I remember talking to one of the doctors about the brain fog. He was asking how it was affecting me and he shared his personal experience of brain fog. You know when he had Long Covid. So, you know you really felt connected with somebody who was genuinely listening to you and trying to do the best for you. (Male, 70-79, White British)*

However, there was also acknowledgement that the support received was clinician-dependent and that it was clear which clinicians were interested in, and are knowledgeable, about Long Covid care.

> *They have been really supportive…but it really depends who you speak to, like you have to speak to someone who actually has an interest in Long Covid and actually wants to learn about it and actually- you know, and that really comes across on conversations and things. (Female, 40-49, White British)*

Participants felt that although their clinicians were supportive and that Long Covid services were good for symptom management, options beyond these were limited.

> *And my experience of dealing with them was really positive. I mean, it’s trying because there aren’t really any treatments for things other than the symptoms that are maybe linked into other things like asthma or whatever. So, you know, it doesn’t fix you necessarily. You get help with managing your symptoms. (Female, 40-49, White)*

Although some participants felt that Long Covid services were ‘innovative’ and demonstrated ‘proper joined-up care’, which could be used as a model for other conditions, not all agreed. Some participants felt that Long Covid care was not set up for the needs of people living with Long Covid. Others felt that learning could be improved, more resources were needed, and better communication would improve Long Covid care. One participant in particular requested an emphasis on improving these services: *Can we just put like, do better, in capitals? (Female, 30-39, White British)*

### 3.3. Theme 2: Disparities when seeking access to Long Covid care

Despite the positive experiences highlighted, some participants felt they experienced unfair treatment when seeking Long Covid care. Gender, race, and health-related discrimination was described. There were also financial barriers to accessing treatment and care.

#### 3.3.1. Testimonial injustice

Testimonial injustice refers to the discriminatory practice whereby an individual’s narrative is dismissed as uncredible due to prejudicial assumptions made about the individual’s characteristics.^31^ Some participants in this study felt their symptoms were dismissed on the basis of their gender or ethnicity, or due to other existing diagnosed health conditions.

##### 3.3.1.1. Gender discrimination as a barrier to accessing care

A number of female participants described feeling discriminated against and dismissed on the basis of their gender when seeking care for symptoms from primary care and/or secondary care services.

> *I did feel like I was being dismissed as sort of a silly woman really. Yes, had it been my husband who was going to see him I think his experience might have been a little bit different. (Female, 40-49, White British)*

Despite feeling that they were treated unfairly because of their gender, women felt they did not have evidence of this behaviour.

> *I had a suspicion that I can’t vouch so whatsoever, but I had a suspicion that if it had been my husband calling not me, he would have been listened to more. Now those are- I can’t back those up. But that is the feeling I had. (Female, 40-49, White other)*

A few female participants felt that women were more likely to have physical symptoms attributed to mental health conditions.

> *I’d say it was just that one doctor really. I don’t know, I feel like as well, women, almost like, things get pinned on mental health, more likely than if you were a man going in. I don’t have any direct proof of that, but that’s just how I felt. He was an older man and just kind of, you’re just- you know, that was the impression I got, that I was just an anxious, young woman [laughs]. (Female, 30-39, White British)*Sometimes gender discrimination was experienced outside of the healthcare system. For instance, a female participant felt her symptoms were dismissed by others when they talked about Long Covid.
>
> *Like, this guy, the other day, he was completely mansplaining how I should feel. He just left me- I was mad. I was angry. I was just like, who are you to tell me how I feel, how I should feel, how I am dealing with my illness?… (Female, 40-49, White)*

##### 3.3.1.2. Racism as a barrier to accessing care

A few participants talked at length about negative experiences within healthcare that they considered to be resulting from racism. One felt that she was treated differently because she was from a Mixed ethic background.

> *They treated my husband differently. I’m half-Asian and he’s white. My name’s [participant’s name] and his is [husband’s name]. So, for me, it was all about my ethnicity. (Female, 50-59, Mixed ethnic background)*

She felt that racism was the main reason for delays to her treatment. And although she did suggest that there might be other reasons for this, she felt that this differential treatment was still ongoing.

Another participant felt that racism and cultural assumptions stood in the way of accessing care.

> *Now in the 80s and 90s when I was growing up a lot of services would assume that because you’re part of an Asian family you’ve got this massive extended family who will help you…I certainly found in terms of my own personal experience that assumption came back in again and they were like, ‘You must have family?’ and I was like, ‘Well actually no. …We haven’t got this set up, we haven’t got this huge community of people helping us.’ and I came up against that. It wasn’t a big thing but it came out quite often and consistently that actually because of who I am, my ethnicity, that I’m going to get all of this help. (Female, 40-49, Asian British)*She also detailed her experience with an Occupational Therapist (OT) who suggested she cut her hair. This suggestion was made to facilitate selfcare and manage fatigue. However, this comment was ignorant to the significance of hair in the participant’s religion, as well as the negative toll this would have on her self-esteem.

One participant described how negative consequences of her inability to speak were compounded by language barriers.

> *For the first year I was mute. So I couldn’t speak and I couldn’t verbalise which was really frustrating for me, I am obviously clearly from a BAME background, my mum ended up being my carer and her English isn’t so good so I had this added barrier which I’ve never really faced before because I’ve been able to advocate for myself and my family where actually, my ethnicity, I was really shocked… just how much my background and the fact that I had a parent who wasn’t able to communicate really changed my outcomes and the way that I was perceived by different services. (Female, 40-49, Asian British)*

Sometimes, participants talked about experiences of racism outside of healthcare. One participant suggested that the COVID-19 pandemic highlighted the extent of racism within the community.

People who frequently experience racial discrimination and prejudice in their everyday lives are likely to be more sensitive to microaggressions experienced when accessing healthcare.

> *And how to live in a world that doesn’t accept my skin colour. Because what COVID did was make racism right there in your face. It’s like living in the 80s again… And being called a [racial slur] every day, sorry about the language but that’s what they referred to me as. So, I’m having to deal with that all again. (Female, 50-59, Mixed ethnic background)*

##### 3.3.1.3. Health-related stigma and discrimination

Participants with existing health conditions – physical and mental - felt that their symptoms were not investigated because of these other conditions.

> *Having multiple health problems I don’t think has helped me because I think initially…a lot of my fatigue and exhaustion and feeling low was put down to bipolar depression. (Female, 40- 49, White British)*
>
> *It was getting the GP to listen because they didn’t seem to want to make a diagnosis of Long Covid. It was, oh, the breathing’s probably down to asthma, or your weight or- I know the weight plays a part, I’m not silly, I do know that and at the moment I’m trying to lose weight… And, yes, it was extremely difficult, not only to get an appointment with the GPs, which is ongoing, but also to get them to actually listen and acknowledge that there was an issue. (Female, 50-59, Asian British)*

This participant had the intersectional dimension of having symptoms attributed to her weight which worked as an additional barrier.

Some participants also talked about how their existing conditions had been exacerbated by their Long Covid symptoms. Those who had previous experience of post-viral illness expressed how they knew what to expect in terms of Long Covid care. In the case of one participant, this meant they were not disappointed with the care they received from the Long Covid service as they acknowledged there is no gold-standard treatment.

> *And I think sometimes people going in are expecting that they’re maybe going to solve everything. And post-viral fatigue illnesses, it doesn’t necessarily work that way and I knew that already. (Female, 40-49, White)*

The stigma was exacerbated if participants has preexisting stigmatised health conditions. One participant, who was diagnosed with myalgic encephalomyelitis (ME) around 11 years prior to her experience with Long Covid, was disheartened by her experience.

> *In fact, I’ve not been told anything new at all during my treatment…it’s actually probably worse than the approach to treatment in the ME Clinic. It’s like there’s all of this information and knowledge that could have been accessed, and utilised, and built upon to help people with Long Covid, and it’s not being used…So, my experience is disappointment and trauma… (Female, 30-39, White British)*

Despite acknowledging that there is ‘*a lot less medical gaslighting going on’* now, she felt that generally Long Covid is a stigmatised condition in society.

> *I was really embarrassed when I was first diagnosed with ME because there’s so much stigma around it. And this has felt quite triggering of that somehow – the guilt, and the shame, and the embarrassment…It feels like it’s happening all over again because Long Covid is being politically brushed under the carpet and not talked about in the news, and so it’s not a thing… And it feels very much like people who still talk about Covid like it’s a serious thing, they’re a joke. (Female, 30-39, White British)*

Similarly, some participants talked about how they felt they were treated differently by some healthcare professionals (HCPs) if they mentioned Long Covid.

> *The minute that you say it’s Long Covid…there just seems to be a barrier wall that they’re not interested…I get so frustrated and I think please just…do something to help but…you can see when you’re explaining it…the look in the doctor’s or consultant’s faces that they just think, you just know you’re not going to get anywhere. (Female, 40-49, White)*

#### 3.3.2. Financial constraints to accessing and receiving care

Some participants experienced financial barriers to accessing treatment and care. This was due to the associated transport costs with attending healthcare appointments as well as the high costs of treatments outside of the NHS which were limited for some.

> *Rheumatology said to take Magnesium which we are already buying…I can only afford to…take it every other day because I want my husband to have the Magnesium too and financially we can’t afford to live as it is… (Female, 50-59, Mixed ethnic background)*

Sometimes financial barriers were heightened by a loss of income and difficulties in obtaining financial support. Ability to apply for financial support, including completing long forms, can also be restricted by chronic illness. Additional barriers to applying for financial support were also experienced by people who are self-employed.

#### 3.3.3. ‘First wavers’: Challenges around accessing care without a documented COVID-19 infection

Nine participants were infected during the first wave of the COVID-19 pandemic in early 2020. This presented its own challenges to obtaining treatment and care for symptoms. During this time, infection control policies and HCP staff shortages meant that some diagnostic tests were delayed. Some participants talked about how an early obstacle to care was that Long Covid was unknown and medical advice was limited. People who contracted COVID-19 early in 2020 were also unable to provide evidence of infection. These participants expressed how this made obtaining referrals from their GPs to Long Covid services more difficult. It also meant they were excluded from some experimental treatment and -as this group of people have been sick for longer- this was regarded as an additional unfairness.

> *I feel that I understand the logic behind it because it creates a study which is more solid than some flaky person who comes along and says, “I’ve had it,” rather than showing them proof… but then at the same time it actually excludes quite a large chunk of people who actually suffered really badly some of us and we are the ones who seem to be stuck and not improving over time. (Female, 50-59, White British)*

To summarise this theme, inequalities in experiences in access to Long Covid care were demonstrated by participants being treated unfairly, in the form of gender discrimination and racism. Layered/compounded stigma was experienced by participants who were intersectionally disadvantaged with preexisting conditions. Participants also explained the financial toll of accessing care and highlighted the challenges of obtaining care without documented evidence of a COVID-19 infection early in the pandemic.

### 3.4. Theme 3: Long Covid and Mental health

As described above, symptoms being attributed to mental health conditions were considered a barrier to care for some participants. However, the theme of mental health extends beyond this and is multifaceted.

#### 3.4.1. Mental health impact of Long Covid

Participants described the impact Long Covid had on their mental health describing anxiety and depression as both symptoms but also as a result of their other symptoms. One participant revealed that her experience had made her feel ‘suicidal’.

> *My memory is not good, and it frustrates me, so I have to- they’ve taught me to write lists and how to break them up into little pieces and do them because I got stressed, and then anxious, then depressed. And fortunately, I eventually realised that I should seek help for that, so I did. I’m now on medication for that. (Female, 50-59, Asian British)*
>
> *And I’m not depressed, I don’t have anxiety, but I definitely have stress-related mood variation, I think, due to my situation. It gets worse when my physical symptoms are worse..(Female, 40- 49, Asian British)*
>
> *My OT has referred me to a health psychologist to try and deal with my- She said, I’m grieving because I’ve lost the person I was before Covid, so I’ve been referred to health psychology for that. (Female, 40-49, White British)*

Other participants talked about how Long Covid symptoms impacted on their self-esteem and had experienced shame, isolation and loneliness, which are examples of internalised stigma.

One participant talked about how her wellbeing has improved since engaging with her Long Covid service and the improvement helps her to manage her physical symptoms.

> *I think talking to the Long Covid clinic that’s where it’s helped me…I don’t feel any different health-wise but psychologically yes, and I think that’s a good step in the right direction…if you are psychologically in your head and your wellbeing is getting better you can cope with things more and I think that’s what has happened because I am coping with things better, I’m dealing with it, I’m understanding it and I am working with it to my advantage so I don’t miss out on things. (Female, 50-59, White British)*

#### 3.4.2. Mental health support available to people with Long Covid

Eight participants reported receiving some form of mental health support, either through primary care or a Long Covid service. Some of these participants found this useful and were appreciative of this support. For others, there were barriers to obtaining the support needed. Psychological support was considered too short-lived:

> *The first thing that came through was the phone therapy…It was kind of helpful, but the problem is that when you’re dealing with something that’s so long-term, having a short-term intervention actually is kind of frustrating because you give your very limited energy to do it and then you don’t have time to get anywhere with it, to really get any benefit from it. Like six 45-minute talking sessions on the phone isn’t really enough to process the trauma of losing your life as you know it, you know what I mean? [laughing]. (Female, 30-39, White British)*

Some felt the waiting times for mental health support were too long and others felt support was non-existent or not appropriate for people with Long Covid.

> *I think…talking to someone who understands…Long COVID, makes a real difference, compared to someone who doesn’t. And like every single [therapist] in the community may not understand that… (Female, 40-49, Indian)*

A few participants felt that their symptoms were only taken seriously once HCPs identified a ‘physical’ cause for symptoms.

> *I would say…being diagnosed with Sjogren’s and then with the myocardial, pericardial helped me to access better care and to be taken more seriously that if I would not have had those conditions. (Female, 40-49, White other)*

Other participants felt that the mental health aspect of Long Covid was not acknowledged and getting mental health referrals was difficult prior to engaging with Long Covid services.

> *It was extremely difficult…to get referrals like to the mental health team. 18 months waiting list and it wasn’t until I…saw the Long Covid clinicians in the clinic that they were able to, via their teams, get me to see someone. (Female, 50-59, Asian British)*

#### 3.4.3. Mental Health Stigma

Some participants suggested that the potential of having their illness wholly attributed to mental health was preventing other people from seeking help for symptoms.

> *I think people are wary because they don’t want to be psychologised into, you know, stuff so obviously they don’t access things because it’s such a diverse condition (Female, 40-49, White British)*

One participant’s feeling of shame related to her mental health had prevented her from reaching out for support.

> *Took me ages to actually tell my GP and actually a couple of close friends…because I felt ashamed, and like a failure that I couldn’t cope with things, and that I should be able to cope…And there are worse things going on in the world and yet here’s me upset, anxious, stressed, depressed for what reason?…So, it took a while for me to feel able, to feel comfortable, to tell anyone and to reach out for care. (Female, 50-59, Asian British)*

### 3.5. Theme 4: Experiences of NHS professionals with Long Covid

Just under the half of the participants worked within the NHS. The following section outlines experiences of barriers and facilitators unique to these participants with some linking to similar concepts emerging from participants from non-NHS backgrounds.

#### 3.5.1. Self-efficacy and self-advocacy as perceived facilitators to accessing care for NHS staff with Long Covid

Participants who worked in healthcare-related roles talked about the ways in which their job facilitated their access to care. Having medical knowledge was considered to be helpful to some of these participants.

> *I think, yes, having that medical knowledge and going in and knowing what to ask for almost, has been the most helpful. (Female, 30-49, White British)*

Sometimes having a medical background was used to evaluate or ‘appraise’ care options. Others were able to monitor their symptoms and take observations at home, using a pulse oximeter, for example, meaning that they were noticing patterns.

People with Long Covid who worked within the NHS felt that they were better equipped to advocate for themselves. Some felt that they had greater access to information and knowledge than the general public.

> *So I should probably mention that I am a doctor, so I have a bit of knowledge about how things work, and if you didn’t have that knowledge, I feel it would be quite difficult. I kind of, went in and knew the right things to ask for, even with the brain fog and everything. (Female, 30-39, White British)*

This included using professional contacts to access Long Covid care. For some, this was considered to be an unfair advantage and there was recognition of the difficulties others might face when navigating the system.

> *I’m also not completely convinced that if I hadn’t happened to have been not just a doctor, but a…doctor with personal contacts at the clinic, I’m not sure that I would have gotten in it really. And I really don’t like having to do that. I would much rather be a proper patient rather than have to mix, sort of, pulling strings essentially to get seen. So, I do really worry about pretty much everybody else. (Female, 40-49, White other)*

Furthermore, some participants who worked within the NHS expressed recognition of what patients experience on the other side of the HCP-patient relationship, including acknowledgement of preconceptions surrounding chronic illness.

Participants who did not work within the NHS talked about having to be ‘pushy’ and having to demand or insist on obtaining care for their symptoms, which suggests that these participants also had the ability to advocate for themselves. However, the perception of self-advocacy and self-efficacy as a facilitator to care among NHS workers differs as NHS workers felt this facilitator originated from their medical knowledge or understanding of the healthcare system that they worked in.

#### 3.5.2. Impact of Long Covid on working within the NHS

A couple of participants felt that NHS workers were at a disadvantage due to their increased risk of contracting COVID-19 and lack of adequate personal protective equipment at the beginning of the pandemic. This was heightened by the experience of seeing people hospitalised with COVID-19.

> *And I think it’s, you know, the frontline saw so many people sick and it’s been really tough for everyone in the pandemic. (Female, 40-49, White British)*

Most of the participants in this study were unable to work due to ill-health at the time of interview, including some participants who worked within healthcare. But despite this, some felt that the NHS, as an institution, had left them to suffer without treatment and without adequate support at work. Sick pay policy was considered unclear or unfair by some.

> *I’m a salaried GP. I’ve found it very difficult that…hospital doctors get paid, they’ve remained on full, or half, whatever their sick pay is, and I haven’t. And no GPs have. (Female, 40-49, Indian)*

Other participants were able to return to work by modifying their usual working tasks.

> *When I went back to work the first time I felt really unsafe to do certain tasks. So, I was on like sort of reduced duties because I wanted to make sure that I wasn’t doing anything that could put anybody in danger. (Female, 40-49, White)*

However, a few participants felt ‘a massive amount of guilt’ taking time off work for their symptoms or not being able to work in the same way as previously.

Sometimes there was inflexibility in terms of adjustments that could be made to working patterns.

> *As an NHS Trust, I think, it’s quite sad because there was probably different options that I could have done but it was like looking at the bigger picture of managing chronic illness and work, you know, but it was just quite black and white so it just was a bit short sighted. (Female, 40- 49, White British)*

A few participants were also concerned about confidentiality related to receiving care within their employment NHS Trust.

> *I didn’t want to get Long Covid care at the hospital I work at because although we have the right to confidentiality…I know they have breached my confidentiality in the past. (Female, 40- 49, Asian British)*

Some participants who did not work within healthcare also talked about the impact of Long Covid on their work. These participants talked about this in relation to not being able to work or having to adjust their working patterns.

#### 3.5.3. Discrimination, stigma and mistreatment from NHS colleagues

Discrimination and stigma were experienced by some NHS workers. Some participants felt their colleagues did not believe them. Despite acknowledgement that Long Covid knowledge was improving, some participants experienced a lack of understanding, care or empathy from colleagues.

> *I had [a] very unpleasant encounter with one of my consultants regarding my hours, my position etc. He was so confrontational and unhappy with me…Saying that how come I can work on one day more hours and not work on other days, and who is checking my hours and kind of threatening if I would work only certain hours I can lose my position etc. (Female, 50- 59, White)*

Other participants expressed being ‘shunned’ or receiving poor treatment from colleagues.

> *I have overheard a couple of conversations between colleagues at work, where they have been like, oh, how many days is she in for this time? Or you know, let’s see how long it lasts, and stuff like that. (Female, 40-49, Asian British)*

One participant felt that she would be seeking alternative employment due to discriminatory experiences. Additionally, one participant felt that her intersectional experiences of feeling discriminated against due to being female and Asian have been deepened as a result of Long Covid.

> *There has always been discrimination, race and sex discrimination but it has been kind of not so overt but once I have got Long Covid, it is like I am the black sheep and I just need to get out. (Female, 40-49, Asian British)*

Two participants who worked outside of healthcare also talked about the lack of support they had received from their employers. Neither of these participants suggested that they had experienced discrimination.

#### 3.5.4. Being at odds with treating clinicians

Sometimes there was disagreement between HCPs with Long Covid and the medical teams responsible for their care, prior to accessing Long Covid services. There was one incident where a participant who worked as an NHS doctor had to ‘agree to differ’ with the opinions of her treating clinicians.

Another participant, who worked in an NHS mental health service, recalled the detrimental impact of a secondary care clinician suggesting a counselling referral for her physical symptoms. This participant talked about being bullied into accepting the referral despite protesting and asserting her knowledge.

To summarise, although some people with Long Covid who worked within healthcare felt that their role facilitated access to care, it was evident that some also experienced difficulties related to employment and quality of care.

## 4. Discussion

Many of the participants recalled positive experiences with primary care and the vast majority of participants stated that the treatment, care and support received improved once their referral to a Long Covid clinic had been fulfilled. NHS Friends and Family Test data support this finding, with 97% reporting a positive experience of Long Covid clinics overall.^32^ These experiences were usually facilitated by positive interactions with clinicians who listened, believed and supported people with Long Covid, despite limits in knowledge and resources.

The data from our interviews also show that people with Long Covid can experience long waits for referrals, diagnostics, and treatment. However, it is important to note that delays following referral to specialist care may not be specific to people with Long Covid. Demand has gone up and long waiting-times are experienced across the healthcare system, with the COVID-19 pandemic exacerbating these issues.^33,34^

For some participants, the intersectional dimensions of peoples’ identities are perceived to influence and shape the experiences of some people accessing care. Unfair treatment was experienced and sometimes perceived as gender and race discrimination, as well as further stigmatisation for people with previous and existing conditions. Financial barriers to accessing treatment were also evident from our analysis, as were difficulties in obtaining care for those who suffered the acute infection in early 2020. Long Covid can impact mental health. Despite this, mental health support and service provision for people with Long Covid were considered to be inadequate and the stigmatised nature of mental health conditions was considered a barrier to seeking support for some participants.

NHS employees felt their care was facilitated by their ability to advocate for themselves as well as their abilities to evaluate their symptoms and care. However, NHS workers expressed how their ability to work was impacted by their Long Covid symptoms, and employment support was sometimes insufficient. We also heard descriptions of perceived discrimination and mistreatment from colleagues as well as experiences of own knowledge being discredited. Although a few participants who did not work within healthcare reflected on the lack of support they received from employers and having to adjust their working patterns, this did not seem to be as common an occurrence as for those working within the NHS. This could be because the majority of non-NHS employees were not in employment at the time of the interview.

Some of our findings were aligned to what other Long Covid studies have found, include findings related to long waits for appointments, referrals, diagnostics and treatment,^25,26,35–37^ experiences of stigma and discrimination,^6,21–23,26,38–40,45^ financial burdens,^35^ difficulties accessing care without confirmation of COVID infection,^37^ the impact of Long Covid on mental health,^35,36,41^ dismissal of symptoms,^23,25,26,35,45^ impact of symptoms on NHS workers ability to fulfil their role and employment support available to NHS workers,^46^ a lack of support from NHS colleagues^46^ and the importance of knowledgeable clinicians.^22,25,37^

Strangl et al.^24^ developed the Health Stigma and Discrimination Framework with the intention of providing a lens for exploration of interactions between health conditions and social location. This study builds on this by demonstrating that Long Covid can be a stigmatised condition in different ways for different population groups. This was experienced by some respondents as gender discrimination, racism and stigma, including internalised stigma, related to co-occurring physical and mental health conditions.

Experiences of stigma and discrimination were reported widely across the sample but manifested in different ways, depending on their individual characteristics. Despite the perception of some that their NHS position worked to facilitate care, it is evident that some challenges experienced when trying to access care were reported to some extent by all our participants.

Stigma and discrimination can prevent people from seeking support for their symptoms. Additionally, stigma and discrimination experienced within the community results in greater sensitivity to experiences, and expectations, of poor treatment within healthcare. Stigma can impact on institutions as well as people.^24^ This can be seen within healthcare, and therefore needs to be acknowledged and tackled at the societal level. The mental health impact of Long Covid also needs to be acknowledged in a way that is destigmatising and does not seek to attribute Long Covid symptoms to a wholly mental health condition.

In our previous work recruiting from communities in England, rather than from health services, we explored barriers and facilitators to accessing care amongst those who do not -yet- have a formal clinical diagnosis of Long Covid.^23^ Similar to the findings from this study where participants described discrimination and disbelief on the basis of their gender or ethnicity, the findings were consistent with the framework of testimonial injustice which is a form of epistemic injustice, or inequality in creating and interpreting knowledge.^31^ Hermeneutical injustice is another form of epistemic injustice where an individual lacks the resources needed to make sense of their lived experience.^31,42^ There was less evidence in this study around hermeneutical injustice as participants mostly had the knowledge needed to consider Long Covid as a cause of their symptoms, however there was inadequate recognition of it in the healthcare system.^45^

Studies exploring the experiences of people with chronic illness have found that epistemic injustice occurs when decisions are made by HCPs to consider some patients as less trustworthy. ^43,44^ Epistemic injustice also occurs when patients lack the ability to express themselves in a way that is understood or considered credible by HCPs.^43,44^ There is evidence that for some people with Long Covid, their experience is that they are not believed or taken seriously, which is defined as epistemic injustice.^23,45^

Long Covid prevalence among people working within healthcare in England is higher than in most other employment sectors.^7^ The reports of the participants within this sample show that some people living with Long Covid and working within healthcare have difficulties fulfilling their NHS role, while also feeling unsupported by the system and their colleagues. The NHS staff survey 2023 found that 30% of NHS employees did not feel valued by their team and 9% had experienced discrimination from colleagues.^47^ This is against the backdrop of a shortage of staff working within the NHS in England.^48^ One of the most commonly cited reasons for leaving the NHS is ill health.^49^ If the wellbeing of staff is not prioritised, this could lead to further exits as demand for healthcare services continues to increase.^48^ The NHS Long Term Workforce Plan acknowledges this and encourages integrated care systems (ICSs), local collaborations including representatives from the NHS, local authorities and voluntary sector, to back development of occupational health and wellbeing programmes. It also suggests fast-tracked access to mental and physical health services for staff as a potential possibility.^48^

As highlighted by others,^22,37^ participants in this study also underlined the importance of positive experiences with knowledgeable clinicians, who have the resources to provide quality care. This further reinforces the need for education and training promoting awareness of Long Covid, as well as sustained dedicated Long Covid services, to ensure that positive interactions with clinicians, who are alert to the symptoms and impact of Long Covid, are a more universal experience. This could also lead to more positive experiences for people living with Long Covid and working within the healthcare system.

### Strengths and weaknesses

People with lived experience of Long Covid were co-investigators on this study and contributed to its design. This study is strengthened by including the experiences of people with Long Covid from three different Long Covid services and the inclusion of people with Long Covid working within healthcare in addition to those from other backgrounds.

Very few people over the age of 60 were recruited to this study. Likewise, the experiences of younger people (<29years) and people from Black or Black British backgrounds are absent from this analysis. The overrepresentation of females in this study can be partially seen as reflective of the gender composition of patients at Long Covid services. In April 2023, 67% of people accessing a Long Covid service in England were female.^12^ However, the number of female participants in this study is still disproportionate to male participants. There is also the possibility that more women self-selected for interview due to a desire to share their experiences related to inequalities. Nonetheless, this does mean that this study may be lacking in terms of experiences of men living with Long Covid. As study interviews were completed online, those without digital literacy or internet access may have been excluded from this study. Future research should focus on recruiting a more diverse sample.

As participants were not asked their motivation for taking part, we do not know why a high proportion of NHS staff expressed interest. Perhaps due to their employment background, they had a greater interest in being interviewed about their healthcare experiences. It is also possible that those who did volunteer to take part did so because they wanted to voice their negative experiences.

As participants self-selected for interview, and the sample is not representative, we cannot make any claims regarding the generalisability of findings. However, this does not minimise the experiences of the people with Long Covid who took part in the interviews. Although the experiences detailed may not be universal, learning from these experiences could ultimately improve circumstances for more service attendees and healthcare staff.

### Main messages

Based on our findings, the main messages we would like to convey to improve Long Covid patient care are:

- Long Covid consultations need to be accessible for all. The location and timing of appointments should be considered. Consultations should be flexible and receptive to the needs of people with Long Covid. These should go at the pace of the patient and be long enough for concerns to be expressed.
- A standardisation of investigation and treatment pathways, which includes provision for employment, benefit and mental health support tailored to people with Long Covid is needed. This requires continued and increased resources for Long Covid services including more staff, more time, and more prioritisation of Long Covid to reduce referral waiting times and move faster with treatment plans.
- Long Covid education for primary and secondary care staff should encourage clinicians to listen and discuss concerns, acknowledge and be openminded about things that might help.
- Long Covid information should be provided within the community to help case finding.
- Primary care teams should be trained to provide support while waiting for referrals.
- Work to mitigate stigma; for example training for healthcare staff which encourages sensitivity to the intersectional barriers that can shape experiences of discrimination.

We propose that addressing these areas is likely to facilitate care and improve the wellbeing of everyone living with Long Covid, including those working within healthcare. These main messages are summarised in figure 1.

**Figure 1.**
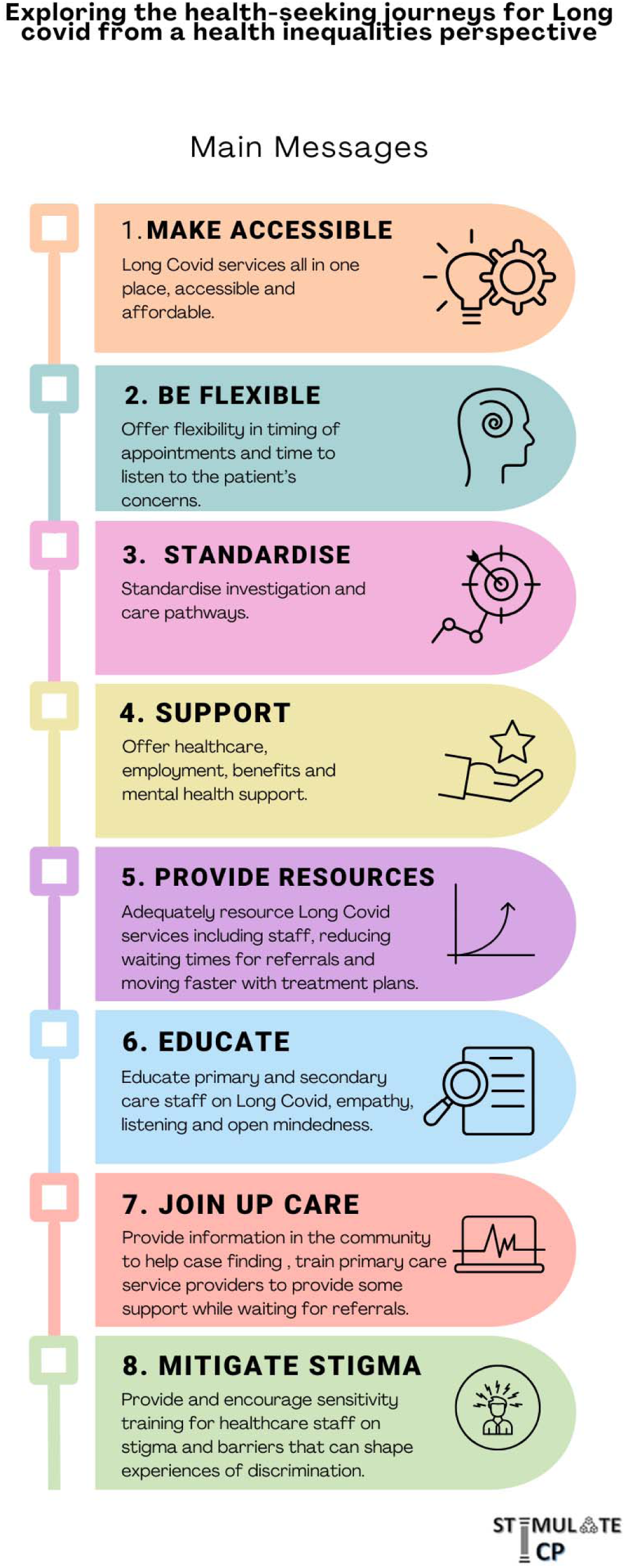
Main messages.

## 5. Conclusion

These findings highlight the difficulties and unfair treatment some people with Long Covid face when accessing treatment and care. Findings also provide further evidence that Long Covid, like some other LTCs, is a stigmatising experience that intersects with the life experiences of different population groups. An intersectional approach, that is mindful of the different ways that stigma and discrimination can impede on experiences and perceptions of care depending on population group, is paramount when exploring lived experience of Long Covid and other chronic conditions. Based on findings from this study, we recommend working towards a more supportive healthcare system that acknowledges and is sensitive to differentials and experiences when accessing care.

## Author Contributions

NAA, MP, MG, GA, AB and JH conceptualised the original idea and obtained funding for the original study as part of STIMULATE-ICP. NAA, MP and DC designed the methodology for this part of the study. DC completed all data collection and led data analysis. MR completed additional coding and contributed towards analysis. DC, MR, MP and NAA developed the thematic framework which forms the basis of this manuscript. DC and NAA drafted the original manuscript. All authors read, reviewed, and approved the final manuscript.

## Acknowledgements

Our sincere thanks to the participants who shared their experiences and contributed to this study.

This research forms part of the STIMULATE-ICP study. An up-to-date version of STIMULATE-ICP Consortium members can be found on https://www.stimulate-icp.org/team. STIMULATE-ICP can be contacted at.

## Ethics

Ethical approval for the project was granted by the Berkshire Research Ethics Committee and the NHS HRA, REC reference: 22/SC/0047.

## Data Availability Statement

The anonymised data that support the findings of this study can be made available on reasonable request from the corresponding authors provided ethics and research governance approvals are obtained.

## Open Access

For the purpose of open access, the author has applied a Creative Commons Attribution (CC BY 4.0) licence to any Author Accepted Manuscript version arising.

## References

1. World Health Organization. A clinical case definition of post COVID-19 condition by a Delphi consensus. Available from: https://www.who.int/publications/i/item/WHO-2019-nCoV-Post_COVID-19_condition-Clinical_case_definition-2021.1 [Accessed 15th February 2024].

2. National Institute for Health and Care Excellence, COVID-19 rapid guideline: managing the long-term effects of COVID-19. Available from: https://www.nice.org.uk/guidance/ng188/resources/covid19-rapid-guideline-managing-the-longterm-effects-of-covid19-pdf-66142028400325 [Accessed 15th February 2024].

3. Woodrow M, Carey C, Ziauddeen N, Thomas R, Akrami A, Lutje V, et al. Systematic Review of the Prevalence of Long COVID. Open Forum Infectious Diseases. 2023;10(7). DOI: 10.1093/ofid/ofad233

4. Ziauddeen N, Gurdasani D, O’Hara ME, Hastie C, Roderick P, Yao G, et al. Characteristics and impact of Long Covid: Findings from an online survey. PLOS ONE. 2022;17(3):e0264331. DOI: 10.1371/journal.pone.0264331

5. Buttery S, Philip KEJ, Williams P, Fallas A, West B, Cumella A, et al. Patient symptoms and experience following COVID-19: results from a UK-wide survey. BMJ Open Respiratory Research. 2021;8:e001075. DOI: 10.1136/bmjresp-2021-001075

6. Ladds E, Rushforth A, Wieringa S, Taylor S, Rayner C, Husain L, et al. Persistent symptoms after Covid-19: qualitative study of 114 “long Covid” patients and draft quality principles for services. BMC Health Services Research. 2020;20(1):1144. DOI: 10.1186/s12913-020-06001-y

7. Office for National Statistics. Self-reported coronavirus (COVID-19) infections and associated symptoms, England and Scotland: November 2023 to March 2024. Available from: https://www.ons.gov.uk/peoplepopulationandcommunity/healthandsocialcare/conditionsanddiseases/articles/selfreportedcoronaviruscovid19infectionsandassociatedsymptomsenglandandscotland/november2023tomarch2024 [accessed 05 July 2024].

8. Graham F, Daily briefing: At least 65 million people have long COVID. Nature. Available from: https://www.nature.com/articles/d41586-023-00114-0 [Accessed 23rd May 2024].

9. Xie Y, Choi T, Al-Aly Z. Postacute Sequelae of SARS-CoV-2 Infection in the Pre-Delta, Delta, and Omicron Eras. New England Journal of Medicine. 2024;391(6):515–25. DOI: doi:10.1056/NEJMoa2403211

10. NHS England. NHS to offer ‘long covid’ sufferers help at specialist centres. Available from: https://www.england.nhs.uk/2020/10/nhs-to-offer-long-covid-help/ [accessed 15th February 2024].

11. NHS England. Commissioning guidance for post-COVID services for adults, children and young people. Available from: https://www.england.nhs.uk/long-read/commissioning-guidance-for-post-covid-services-for-adults-children-and-young-people/ [accessed 15th February 2024].

12. NHS England. Statistics: COVID-19 Post-COVID Assessment Service: 10 April 2023 - 07 May 2023. Available from: https://www.england.nhs.uk/statistics/statistical-work-areas/covid-19-post-covid-assessment-service/ [accessed 02 August 2024]

13. English indices of deprivation 2019. Available from: https://www.gov.uk/government/statistics/english-indices-of-deprivation-2019 [accessed 02 August 2024]

14. The King’s Fund. What are health inequalities? Available from: https://www.kingsfund.org.uk/insight-and-analysis/long-reads/what-are-health-inequalities [accessed 15th February 2024].

15. Smyth N, Ridge D, Kingstone T, Gopal DP, Alwan N, Wright A, et al. People from ethnic minorities seeking help for Long Covid: a qualitative study. British Journal of General Practice. 2024:BJGP.2023.0631. DOI: 10.3399/BJGP.2023.0631

16. Heightman M, Prashar J, Hillman TE, Marks M, Livingston R, et al Post-COVID-19 assessment in a specialist clinical service: a 12-month, single-centre, prospective study in 1325 individuals BMJ Open Respiratory Research 2021;8:e001041. DOI: 10.1136/bmjresp-2021-001041.

17. Dean E. What happens inside a long COVID clinic? BMJ 2023; DOI: 10.1136/bmj.p1791.

18. Collins PH. Intersectionality’s Definitional Dilemmas. Annual Review of Sociology. 2015;41(1):1–20. DOI: 10.1146/annurev-soc-073014-112142.

19. Atewologun D. Intersectionality Theory and Practice. Oxford Research Encyclopedia of Business and Management. 2018; published online Aug 28. DOI:10.1093/acrefore/9780190224851.013.48.

20. Kapilashrami A, Hankivsky O. Intersectionality and why it matters to global health. The Lancet. 2018;391(10140):2589–91. DOI: 10.1016/S0140-6736(18)31431-4

21. Pantelic M, Ziauddeen N, Boyes M, O’Hara ME, Hastie C, Alwan NA. Long Covid stigma: Estimating burden and validating scale in a UK-based sample. PLOS ONE. 2022;17(11):e0277317. DOI: 10.1371/journal.pone.0277317

22. Kingstone T, Taylor AK, O’Donnell CA, Atherton H, Blane DN, Chew-Graham CA. Finding the “right” GP: a qualitative study of the experiences of people with long-COVID. BJGP Open 2020; 4:bjgpopen20X101143. DOI: 10.3399/bjgpopen20X101143

23. Clutterbuck D, Ramasawmy M, Pantelic M, Hayer J, Begum F, Faghy M, et al. Barriers to healthcare access and experiences of stigma: Findings from a coproduced Long Covid case-finding study. Health Expectations. 2024;27(2):e14037. DOI: 10.1111/hex.14037

24. Stangl AL, Earnshaw VA, Logie CH, van Brakel W, C. Simbayi L, Barré I, et al. The Health Stigma and Discrimination Framework: a global, crosscutting framework to inform research, intervention development, and policy on health-related stigmas. BMC Medicine. 2019;17(1):31. DOI: 10.1186/s12916-019-1271-3

25. Sunkersing D, Ramasawmy M, Alwan NA, Clutterbuck D, Mu Y, Horstmanshof K, et al. What is current care for people with Long COVID in England? A qualitative interview study. BMJ Open. 2024;14(5):e080967. DOI: 10.1136/bmjopen-2023-080967

26. Mullard J, Mir G, Herbert C, Evans S, Locomotion Consortium. ‘You’re just a Guinea pig’: Exploring the barriers and impacts of living with long COVID-19: A view from the undiagnosed. Sociology of Health & Illness. 2024. DOI: 10.1111/1467-9566.13795

27. Ramasawmy M, Mu Y, Clutterbuck D, Pantelic M, Lip GYH, van der Feltz-Cornelis CM. et al. STIMULATE-ICP-CAREINEQUAL - Defining usual care and examining inequalities in Long Covid support: protocol for a mixed-methods study (part of STIMULATE-ICP: Symptoms, Trajectory, Inequalities and Management: Understanding Long-COVID to Address and Transform Existing Integrated Care Pathways). PLOS ONE.2022;17(8):e0271978. DOI: 10.1371/journal.pone.0271978

28. Braun V, Clarke V. Thematic analysis. In Cooper H, Camic PM, Long DL, Panter AT, Rindskopf D, Sher, KJ. (Eds.) APA handbook of research methods in psychology, Vol. 2. Research designs: Quantitative, qualitative, neuropsychological, and biological. American Psychological Association; 2012. p.57–71.

29. Braun V, Clarke V. Using thematic analysis in psychology. Qualitative Research in Psychology. 2006;3(2):77–101. DOI: 10.1191/1478088706qp063oa

30. Lumivero. NVivo (Version 13, 2020 R1). 2020.

31. Fricker M. Epistemic Injustice: Power and the Ethics of Knowing: Oxford University Press; 2007. Available from: 10.1093/acprof:oso/9780198237907.001.0001.

32. NHS England. Friends and Family Test (FFT) data collection overview – August 2024. Available from: https://www.england.nhs.uk/publication/friends-and-family-test-data-august-2024/#heading-1 [accessed 8th November 2024]

33. British Medical Association. NHS backlog data analysis. Available from: https://www.bma.org.uk/advice-and-support/nhs-delivery-and-workforce/pressures/nhs-backlog-data-analysis [accessed 02 August 2024]

34. British Medical Association. An NHS under pressure. Available from: https://www.bma.org.uk/advice-and-support/nhs-delivery-and-workforce/pressures/an-nhs-under-pressure [Accessed 02 August 2024]

35. Owen R, Ashton RE, Skipper L, Phillips BE, Yates J, Thomas C, et al. Long COVID quality of life and healthcare experiences in the UK: a mixed method online survey. Quality of Life Research. 2023. DOI: 10.1007/s11136-023-03513-y

36. Callum T, Mark AF, Rebecca O, James Y, Francesco F, Tom B, et al. Lived experience of patients with Long COVID: a qualitative study in the UK. BMJ Open. 2023;13(4):e068481. DOI: 10.1136/bmjopen-2022-068481

37. Turk F, Sweetman J, Chew-Graham CA, Gabbay M, Shepherd J, van der Feltz-Cornelis C, et al. Accessing care for Long Covid from the perspectives of patients and healthcare practitioners: A qualitative study. Health Expectations. 2024;27(2):e14008. DOI: 10.1111/hex.14008

38. Baz SA, Fang C, Carpentieri JD, Sheard L. ‘I don’t know what to do or where to go’. Experiences of accessing healthcare support from the perspectives of people living with Long Covid and healthcare professionals: a qualitative study in Bradford, UK. Health Expect. 2023; 26: 542–554. DOI: 10.1111/hex.13687

39. Macpherson K, Cooper K, Harbour J, Mahal D, Miller C, Nairn M. Experiences of living with long COVID and of accessing healthcare services: a qualitative systematic review. BMJ Open 2022;12:e050979. DOI: 10.1136/bmjopen-2021-050979.

40. Rushforth A, Ladds E, Wieringa S, Taylor S, Husain L, Greenhalgh T. Long Covid – The illness narratives. Social Science & Medicine. 2021;286:114326. DOI: 10.1016/j.socscimed.2021.114326

41. Houben-Wilke S, Goërtz YMJ, Delbressine JM, Vaes AW, Meys R, Machado FVC, et al. The Impact of Long COVID-19 on Mental Health: Observational 6-Month Follow-Up Study. JMIR Ment Health. 2022;9(2):e33704. DOI: 10.2196/33704

42. Bhakuni H, Abimbola S. Epistemic injustice in academic global health. The Lancet Global Health. 2021;9(10):e1465–e70. DOI: 10.1016/S2214-109X(21)00301-6

43. Carel H, Kidd IJ. Epistemic injustice in healthcare: a philosophial analysis. Medicine, Health Care and Philosophy. 2014;17(4):529–40. DOI: 10.1007/s11019-014-9560-2

44. Buchman DZ, Ho A, Goldberg DS. Investigating Trust, Expertise, and Epistemic Injustice in Chronic Pain. Journal of Bioethical Inquiry. 2017;14(1):31–42. DOI: 10.1007/s11673-016-9761-x

45. Ireson, J, Taylor, A, Richardson, E, Greenfield, B, Jones, G. Exploring invisibility and epistemic injustice in Long Covid—a citizen science qualitative analysis of patient stories from an online Covid community. Health Expect. 2022; 25: 1753–1765.DOI: 10.1111/hex.13518

46. Torrance N, MacIver E, Adams NN, Skåtun D, Scott N, Kennedy C, et al. Lived experience of work and long COVID in healthcare staff. Occupational Medicine. 2024;74(1):78–85.

47. NHS England. NHS Staff Survey 2023. Available from: https://www.nhsstaffsurveys.com/results/national-results/ [Accessed 8th November 2024]

48. NHS. NHS Long Term Workforce Plan. June 2023. Available from: https://www.england.nhs.uk/wp-content/uploads/2023/06/nhs-long-term-workforce-plan-v1.2.pdf [accessed 23rd May 2024].

49. Palmer, B. The long goodbye? Exploring rates of staff leaving the NHS and social care. The Nuffield Trust. Available from: https://www.nuffieldtrust.org.uk/resource/the-long-goodbye-exploring-rates-of-staff-leaving-the-nhs-and-social-care [accessed 23rd May 2024].

